# Lipidomic Profiling Unveils Sex Differences in Diabetes Risk: Implications for Precision Medicine

**DOI:** 10.1101/2023.05.06.23289612

**Authors:** Ana F. Pina, Maria João Meneses, Fabrizia Carli, Bárbara Patrício, Rogério T. Ribeiro, Rita S. Patarrão, Luís Gardete-Correia, Rui Duarte, José M. Boavida, João F. Raposo, Amalia Gastaldelli, Maria Paula Macedo

## Abstract

Type 2 diabetes (T2D) is a multifactorial condition whose greatest impact comes from its complications. Not only impaired glucose homeostasis, but also lipid alterations have a relevant role, with insulin derived mechanisms behind this milieu, i.e., glycemia and lipidemia. Thus, we hypothesized that a) distinct glucose and lipid profiles and b) sex differences, particularly in lipids patterns, may be used to identify subjects at higher risk to develop T2D.

The PREVADIAB2 study evaluated metabolic alterations after 5 years in subjects without T2D when participating to PREVADIAB1. Herein, 953 subjects from the PREVADIAB2 cohort were stratified using a hierarchical clustering algorithm, informed by HOMA-IR, IGI, _f_ISR and _f_IC. The resulting clusters were profiled and the lipidome of a subset (n=488) was assessed by LC/MS-QTOF.

We identified four clusters, named according to their main metabolic features: Liver Sensitive (LS); Pancreas Glucose Sensitive (PGS); Insulin Deficient (ID); and Insulin Resistant (IR). These cluster metabolic patterns were similar between sexes. However, men and women had distinct parameters cut-offs and lipidomic profiles. Overall, women presented higher long chain ceramides. Nonetheless, men had higher ceramide to sphingomyelin ratio and higher lysophosphatidylcholine to phosphatidylcholine ratio. For both genders, the LS cluster had the most advantageous lipid profile, whereas the other clusters presented lipid specificities towards dysmetabolism. This work shows that clustering individuals by distinct insulin-related metabolic features and sex identifies different phenotypes with distinct lipidome profile, thus demonstrating the importance of placing diabetes in a broader context of metabolism beyond glucose.

## Introduction

Type 2 diabetes (T2D) is a chronic complex disease, diagnosed by elevated fasting and/or postprandial glycemia or HbA1c. The management of T2D requires multifactorial behavioral and pharmacological treatments, not only to prevent or delay complications, but also to maintain quality of life (1). Definitely, there are other known risk factors for diabetes complications besides hyperglycemia, e.g., dyslipidemia and hypertension (2). In current T2D diagnosis criteria only glycemia is considered (3); however, diabetes develops because of insulin altered secretion, action or metabolism (4-7) and insulin also acts on lipid metabolism (8), which is in turn fully interconnected with glucose metabolism (9).

Diabetes heterogeneity is fully recognized (10), but difficult to tackle. With the use of data-driven cluster analysis (based on glutamate decarboxylase antibodies (GADA), age at diagnosis, BMI, HbA1c, HOMA-IR and HOMA-B) in patients with newly diagnosed diabetes Ahlqvist et al. identified five replicable clusters of patients with diabetes, which had significantly different characteristics and risk of complications (11). Given the high heterogeneity it is important to view diabetes in a broader perspective that encompasses the patho/physiological insulin mechanisms and their consequent metabolites, glycemia and others, as is the case for lipids. Together, a holistic perspective might aid in discriminating diabetes subtypes, how they relate to sex difference and, more importantly, might explain the appearance of complications in some subjects and not in others. However, only few studies have analyzed clusters of subjects with or at risk of T2D including lipidomics analysis (12; 13).

Lipids are a broad and heterogeneous group of molecules, both in structure and in their actions. They encompass 1) fatty acyls (FA); 2) glycerolipids (GL); 3) glycerophospholipids (GP); 4) sphingolipids (SP); 5) sterol lipids (ST); 6) prenol lipids (PR); 7) saccharolipids (SL) and 8) polyketides. They serve mainly energy storage, cellular structure, and intra- and intercellular messengers, amongst others (14). Therefore, lipids metabolism alterations will impact widely in our organism.

In recent years, lipidomic analysis has been developed and used to tackle dysmetabolic conditions (15). Furthermore, it allowed the identification of specific lipids species altered in diabetes (16; 17), diabetes’s related conditions (e.g., NAFLD) (18), and complications (19), and that importantly can be used as biomarkers, including DAGs and TGs (a GLs), ceramides (CERs) and sphingomyelins (SMs) (a SP’s species), phosphatidylcholine (in GP lipids), and others.

Notably, lipidomic profiles were additionally found to be distinct by sex and age in healthy individuals (20; 21), thus suggesting that it might also be the case in unhealthy conditions as in diabetes or in its progression status and might contribute to different sex risk for diabetes complications.

In this work, we hypothesize that by clustering a population regarding a few well-known diabetes pathophysiologic mechanisms, we can define distinct glycemic patterns along with lipidomic profiles. We further hypothesize that there are sex differences specifically in lipids patterns. These patterns can support subgroups’ differentiation as well as explain distinct potential complications. We aim to assess lipids and glycemia profiles of subjects homogeneous in diabetes pathophysiology.

## Material And Methods

### Population

The study population comprises the participants of PREVADIAB2, a follow-up of the first diabetes prevalence study performed in Portugal (PREVADIAB1) (22; 23). In brief, between 2008 and 2009, 5167 subjects attending the national healthcare system across the country were recruited and screened for diabetes. In 2014, we randomly selected 1084 subjects without diabetes in PREVADIAB1. A letter was sent to all participants, that voluntarily agreed to participate in PREVADIAB2 and provided written informed consent. Ethical permits to conduct this study were obtained from the Ethics Committee of the *Associação Protectora dos Diabéticos de Portugal* (APDP). The study protocol followed the Declaration of Helsinki and was approved by the *Autoridade Nacional de Protecção de Dados* (3228/2013).

In the present study, only individuals with complete profiles regarding input parameters for the cluster analysis described below were included. Subjects that had medical treated diabetes were excluded (Supplementary Figure 1). For the retrospective analysis of glycemia progression, subjects that had incomplete data were also excluded from the analysis.

### Clinical and Biochemical Parameters

Evaluation of the participants of PREVADIAB2 was previously described (22). In summary, subjects were evaluated regarding their height and weight. Additionally, they underwent a standardized 75 g oral glucose tolerance test (OGTT), and glucose, insulin, C-peptide and free fatty acids blood levels were measured at 0, 30 and 120 min of the OGTT. The diabetes status of each participant was determined using the WHO criteria for increased risk of (pre)diabetes (24). Furthermore, serum samples were used to analyze glycated hemoglobin and a untargeted lipidomic analysis was performed at fast, as described below.

### Lipidomic analysis

Serum lipidome was analyzed using an ultra-high performance liquid chromatography-quadrupole time-of-flight mass spectrometry system (UHPLC-QTOF; Agilent Technologies). Besides triglycerides, the analysis covered most of the major molecular lipids including ceramides, sphingomyelins (SM), phosphatidylcholines (PC), and lysophosphatidylcholines (LPC).

Briefly, 10 μl of serum were mixed with 10 μl of an internal standards mixture and 150 μl of cold methanol and centrifuged at 14000 rpm for 20 minutes. The internal standard solution contained the following compounds: N-heptadecanoyl-D-erythro-sphingosylphosphorylcholine (SM(d18:1/17:0)), N-heptadecanoyl-D-erythro-sphingosine (Cer(d18:1/17:0)), 1,2-diheptadecanoyl-sn-glycero-3-phosphocholine (PC(17:0/17:0)) and 1-heptadecanoyl-2-hydroxy-sn-glycero-3-phosphocholine (LPC(17:0)) purchased from Avanti Polar Lipids Inc. and tripentadecanoin (TG(15:0/15:0/15:0)) purchased from Larodan AB.

After centrifugation, the methanol supernatant was transferred to the vials for instrumental analysis and 1 μl was injected in UHPLC-QTOF system from Agilent Technologies, that combining a 1290 Infinity LC system and a 6545 quadrupole time-of-flight mass spectrometer (QTOF), interfaced with a dual jet stream electrospray (dual ESI) ion source. Lipids were separated by ZORBAX Eclipse Plus C18 2.1×100mm 1.8 μm column (Agilent, Santa Clara CA) and mobile phases were water with 0.1% formic acid (mobile phase A) and isopropanol:acetonitrile (1:1, v:v) with 0.1% formic acid (mobile phase B). Acquisition was in positive electrospray ionization mode. Profinder MassHunter B.08.00 software (Agilent Technologies) was used for analysis of data acquisition.

### Biochemical and Metabolic Indexes

Insulin secretion rate at fast (_f_ISR) and insulinogenic index (IGI) (25) were used to evaluate insulin secretion. _f_ISR was estimated from insulin and C-peptide values as previously reported using a two compartmental model with population parameters (26). Insulin clearance (L.min^-1^) was calculated from the ratio of peripheral insulin disposal rates and circulating insulin concentrations as previously (26). The suppression of insulin clearance during OGTT (26) was presented as the slope of insulin clearance from 0 min to 30 min (Δ(0–30)IC), and additionally, by the value of insulin clearance at 120 min (IC 120 min). We used the HOMA-IR2 (HOMA calculator) to evaluate the insulin resistance at 0 min. Also, organ-specific insulin resistance indexes were assessed: hepatic-IR (27) as AUC(0–30)Insulin × AUC(0–30)Glucose. Adipo-IR was derived as FFA_0min_ × Insulin_0min_ (28). To evaluate the pancreatic function we considered the OGTT Disposition Index (DI) (29). NAFLD-FLS, a surrogate index was of NAFLD, calculated as −2.889 + 1.179 × IDF metabolic syndrome (yes = 1/no = 0) + 0.454 × type2diabetes (yes = 2/no = 0) + 0.145 × Insulin_0min_ + 0.038 × AST – 0.936 × AST/ALT (30) was used for the detection of NAFLD (31).

### Cluster Analysis

The cluster analysis was done with the agglomerative hierarchical clustering algorithm with Ward method, by using the stats::hclust in R. Considering the sample size, hierarchical cluster algorithm is computational efficient and furthermore allows to explore the dendrogram at different levels (32). The optimal number of clusters was identified by the majority rule, using NbClust package in R (33).

As input variables of the clustering algorithm we chose: _f_ISR and IGI to estimate insulin secretion at fast and during the first phase of the OGTT, respectively; _f_IC to measure insulin clearance at fast; and HOMA-IR as a surrogate index of the overall insulin resistance.

During the preprocessing phase severe outliers were excluded using a multidimensional approach and data was centered and scaled.

Lipid species and other parameters besides the ones informing the cluster analysis were used to profile the clusters namely glucose levels, surrogate indexes of fatty liver and organ-specific insulin resistance. We excluded from the analyses PC C36:6, as it had a high number of outliers and few other outlier samples identified with PCA (Supplementary Figure 2). After preprocessing, we analyzed 103 lipid species in 273 women and 215 men. Lipidomic-clusters association was assessed adjusting for age, BMI, and glycemia subgroups, as these factors could impact on the analysis.

To assess if the clusters’ profiling pattern was stable across glycemia subgroups, we performed a sensitivity analysis where the clusters were profiled individually: subjects with normoglycemia and dysglycemia.

Except for the lipidomic analysis, comparison between groups means (clusters and sex) was performed with Kruskall-Wallis and Wilcoxon-Mann-Whitney test for continuous variables. Chi-square test was used for the comparison of categorical variables. Bonferroni correction was used to multicomparison, when applicable.

Lipidomic data analysis was performed with lipidr package in R (34) after being normalized and log2transformed. Outliers were assessed and filtered using a multidimensional evaluation with PCA (Supplementary Figure 2). Lipids-clusters association was assessed with ANOVA, adjusted to age, BMI and glycemia subgroups: normoglycemia; dysglycemia including prediabetes and diabetes. Benjamin-Hochberg (BH) was used to adjust for multiple comparison, as implemented in the used package.

## Results

### Population characterization and overall sex diferences

We analyzed 953 subjects from the PREVADIAB2 cohort, with a mean age of 61 ± 13; 60% were women. The sample included subjects previously enrolled in PREVADIAB1 study that had no diabetes at inclusion; at screening for PREVADIAB2 subjects resulted within all classes of dysglycemia (70% with normoglycemia, 22% with prediabetes, and 5% with diabetes) (Supplementary Table 1).

Women and men had differences in three of the four parameters used to inform the cluster analysis, as well as in some parameters used only for profiling the clusters (Supplementary Table 1). Specifically, when compared to men, women had higher BMI and higher IGI despite having lower insulin secretion at fasting. Additionally, women had tendentially higher hepatic-IR than men, and lower ISI_0-120_. Nonetheless, there were no differences regarding NAFLD-FLS, adipo-IR, or HOMA-IR (Supplementary Table 1). Consistently, women had lower insulin clearance than men. Finally, women had lower glycemia values at fast and at 30 minutes of the OGTT but higher levels at 120 minutes of the OGTT (Supplementary Table 1).

Women and men showed distinct lipidomic profiles overall. In women, SM species were higher than in men. CER to SM ratio, which might be associated with insulin secretion defects (35; 36), were higher in men (0.90 ± 0.18 vs. 1.04 ± 0.21 for women and men respectively, p<0.001).

Additionally, LPCs and LPC to PC ratio, associated with increased risk of an inflammatory profile (37), were higher in men than in women (4.12 ± 4.6 vs. 1.4 ± 0.6, p<0.001). Women had higher levels of long-chain (C14:0-C20:0) and very-long-chain dihydroceramides species (C22:0-C25:0) than men, which are related with the *de novo* ceramide synthesis (5.3 ± 1.9 vs 4.7 ± 1.8, p<0.001). In contrast, men had higher very long chain and ultralong chain (>C25:0) ceramide species (10.2 ± 3.4 vs 11.6 ± 4.0, p<0.001).

### Cluster analysis

We sought to validate in the PREVADIAB2 cohort if men and women have the same cluster centroids and possible differences in their lipidomic profile.

The cluster analysis workflow is represented in Supplementary Figure 1. We stratified the population regarding three mechanisms related to diabetes pathophysiology, namely insulin secretion (_f_ISR and IGI to estimate insulin secretion at fast and during the first phase of the OGTT), resistance (HOMA-IR), and fasting insulin clearance (_f_IC), as input variables of the clustering algorithm. First we investigate differences among cluster in the whole population and then for women and men separately.

In the cluster analysis performed with the overall population, the best number of clusters was four, that were named based on their metabolic profiles (Supplementary Table 2). The liver-sensitive (LS) cluster is characterized by having the highest IC and the lowest hepatic-IR. The pancreas glucose sensitive (PGS) cluster had the highest DI and IGI. Nevertheless, the PGS cluster had higher HOMA-IR and suppressed IC compared with the LS cluster. Insulin deficient (ID) cluster had low _f_ISR and IGI, although it had higher HOMA-IR than the LS cluster. Insulin resistant (IR) cluster had suppressed IC, the highest HOMA-IR, and consistently the highest _f_ISR but not IGI. When reported by sex, we found that both men and women clusters had similar patterns to the overall population. Nevertheless, they had different cluster centroids (Supplementary Table 2). Thus, both sex could have different cut-off levels for the informing parameters, and therefore we proceeded with a separate cluster analysis for each sex. Additionally, variable importance order was distinct when assessed for the cluster analysis with the overall population and of note for both sex separately, supporting a distinct metabolism pattern (Table 1). Interestingly, HOMA-IR was one of the most important variables in women, whereas for men was the least important.

**Table 1.** Variable importance for cluster analysis. Variable importance in projection (VIP) values of the parameters informing the cluster analysis: of the overall population as well as for both genders separately. IC – Insulin clearance; IGI – Insulinogenic index; ISR – Insulin secretion rate; f denotes fast.

| VIP all | VIP women | VIP men |
| --- | --- | --- |
| IGI<br>(1.1) | fIC<br>(1.1) | fIC<br>(1.2) |
| fIC<br>(1.0) | HOMA-IR<br>(1.1) | IGI<br>(1.0) |
| HOMA-IR<br>(1.0) | IGI<br>(1.0) | fISR<br>(1.0) |
| fISR<br>(0.9) | fISR<br>(0.8) | HOMA-IR<br>(0.8) |

Clusters showed distinct risks for hyperglycemia in both sex. PGS cluster had the lowest risk of having dysglycemia, while the IR cluster had the highest (Figure 1A). A 5-year retrospective analysis of glycemic class evolution was performed (Figure 1B), taking into account that all individuals had no diabetes 5 years before this study. Overall, the incidence of diabetes is higher in 3 clusters: LS, ID and IR. Regarding prediabetes, the highest incidence is in ID and IR clusters. Besides, the latest have higher incidence of prediabetes and diabetes in women compared to men. PGS cluster had the highest proportion of subjects that remained normoglycemic, whereas the IR cluster had the highest proportion of progressors, either to PD or diabetes, namely in women. Nonetheless, almost 25% of subjects in the LS cluster had progressed from normoglycemia to prediabetes or diabetes.

**Figure 1.**
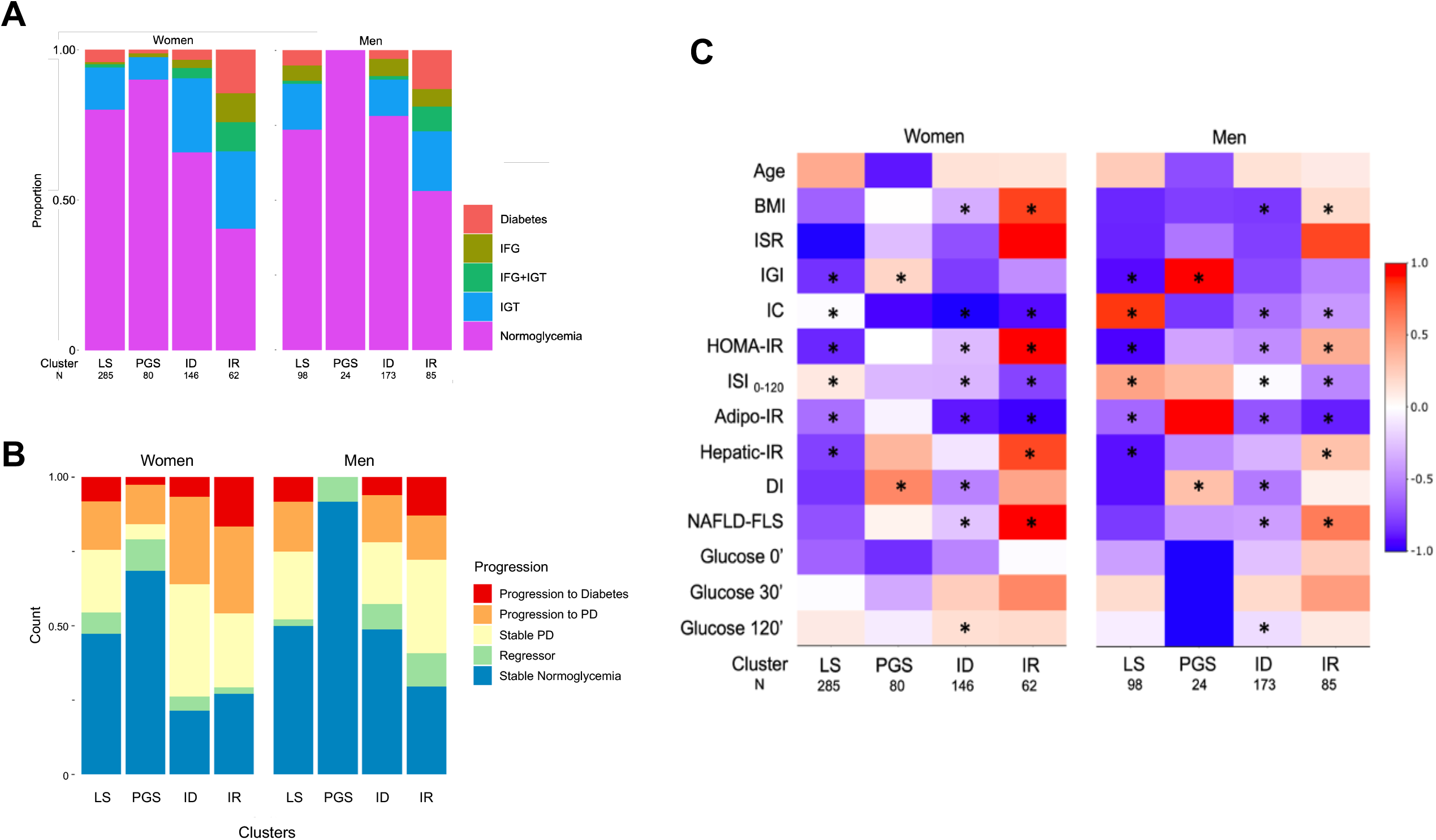
**A** – Proportion of glycemia classes in the clusters by gender. **B** - Retrospective evaluation of glycemia evolution within the clusters, by gender. Proportion of individuals that have progressed, regressed or maintained their glycemic levels in a 5-year period. **C** - Cluster’s profile by gender. Heatmap representing the scaled mean of each parameter by gender. Cluster Names: liver-sensitive (LS); pancreas glucose sensitive (PGS); insulin deficient (ID); insulin resistant (IR).

Cluster’s profiles for women and men are represented in Figure 1C. Men from the LS cluster had higher IC and _f_ISR and lower IGI and insulin-resistance indexes (overall and organ-specific) than women. However, in the PGS cluster, men had higher IGI and DI. Men from the ID cluster had lower _f_ISR than women and lower IR indexes and NAFLD-FLS and higher IC and DI. Additionally, ID men had lower BMI than ID women. Finally, compared to women from the IR cluster, men also had lower insulin-resistance indexes, BMI, NAFLD-FLS, and higher IC. However, they did not differ in insulin secretion indexes - _f_ISR and IGI.

### Clusters lipidomic signatures

Genders showed distinct lipidomic profiles throughout the clusters (Supplementary Figure 3 and 4). From the 103 evaluated lipid species, 79 were significantly associated with the clusters: 42 were associated with both genders; 23 were associated only with women clusters; 14 lipid species were only associated with men clusters (Supplementary Table 3).

Figure 2 reports differences in the lipidomic profile in the four clusters in men and women. Figure 2G represents the clusters profiling for the lipid species that were found to be significantly associated with clusters in women and/or in men (p<0.05, BH correction for multiple comparison). Interestingly, both genders showed clusters with high levels of ceramides, albeit for distinct ceramides species.

**Figure 2.**
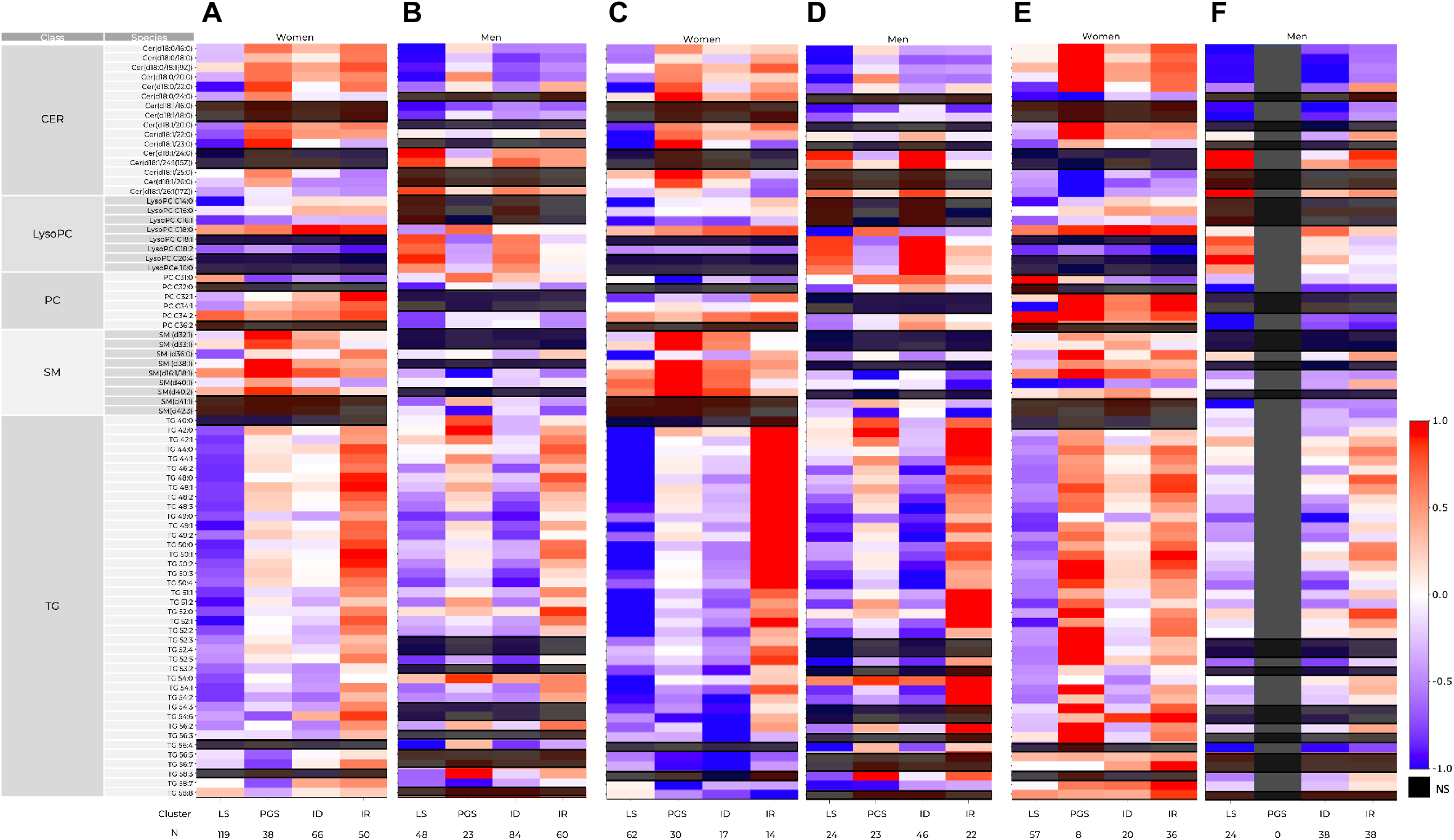
Lipidomics cluster’s profiling. Heatmap representing the scaled mean of each parameter by gender. **A-B** overall population, **C-D** normoglycemia, **E-F** dysglycemia (PD and Diabetes). The analysis was done individually for each gender. From the 103 lipid species 23 were associated with women’s clusters, 14 with men’s clusters and 42 associated with both gender’s clusters. CER – ceramides; ID – insulin deficient; IR – insulin resistant, LS – liver sensitive; LysoPC – lysophosphatidylcholine; NS – non-significant; PC – phosphatidylcholine; SM - sphingomyelin; PGS – pancreas glucose sensitive; TG – triglycerides.

#### Liver Sensitive Cluster (LS)

Women presented lower overall measured CER (Cer(d18:0/16:0), Cer(d18:0/18:0), Cer(d18:0/20:0), Cer(d18:0/22:0)) and TG species, namely TG48:2. Similarly, men had low TG (TG48:2) and medium-chain ceramides (Cer(d18:0/16:0), Cer(d18:0/18:0), Cer(d18:0/20:0), Cer(d18:0/22:0)) compared to the remaining clusters. Nevertheless, they had high levels of long and very-long-chain dihydroceramides (Cer(d18:1/24:0), Cer(d18:1/24:1), Cer(d18:1/26:1)). Interestingly, men also had high LPC18:2 but low LPC18:0.

#### Pancreas Glucose Sensitive Cluster (PGS)

Subjects from the PGS cluster had higher saturated ceramides ((Cer(d18:0/16:0), Cer(d18:0/18:0), Cer(d18:0/20:0), Cer(d18:0/22:0)) and TG levels than LS cluster, particularly TG48:2, a *de novo* lipogenesis marker. This group also presented high levels of SM, particularly in women (SM(d32:1), SM(d33:1), SM(d38:1), SM(d16:1/18:1), SM(d40:1), SM(d40:2)). Finally, this PGS cluster had low LPC14:0, LPC16:1 and LPC18:2. Nevertheless LPC16:0 and LPC18:0 were higher which notably have been associated with decreased T2D risk (38).

#### Insulin Deficient Cluster (ID)

In men, the ID cluster lipidomic profile resembled the LS cluster profile. However, men in ID presented the lowest levels of some TG species (e.g., TG49:0, TG50:4). This result was not evident in women, that have higher insulin resistance and more suppressed insulin clearance than men.

#### Insulin Resistant Cluster (IR)

IR cluster had the most altered metabolic profile regarding glucose and lipid metabolism, in both genders (Figure 1C). Indeed, this cluster present the highest levels of TGs (e.g., TG48:0, TG49:0, TG50:0), specially in women with normoglycemia. Still, some deleterious lipid species showed higher levels in women than in men (e.g., TGs and Cers), suggesting that this population can have a higher risk of cardiometabolic complications.

### Sensitivity analysis

The patterns within each cluster were the same, independently of the glycemic class. Nonetheless, there were few differences in the centroids of the clusters when the subgroups were compared, namely regarding IGI (Supplementary Table 4; Supplementary Figure 5).

## Discussion

This study aimed to evaluate how the pathophysiological mechanisms of diabetes are associated with distinct glucose and lipid profiles and if sex differences, particularly in lipids patterns, should be considered to identify subjects at higher risk to develop T2D.

We performed a hierarchical cluster analysis based on three critical mechanisms in diabetes pathophysiology: insulin secretion, clearance and resistance in a population of 953 subjects (60% women) from the PREVADIAB2 cohort, previously studied in the PREVADIAB1 study 5 years before when the individuals did not had diabetes.

We found 4 clusters that were named according to their metabolic characteristics: LS, PGS, ID, and IR. Consistent with previous work, including deeper profiling, the clusters demonstrated a difference in hyperglycemia risk (lowest in PGS and highest in IR). When we performed the separate analysis by sex, although the metabolic pattern of the cluster remained, the centroids were different. Importantly, our results support that distinct patterns of metabolism, in which alterations in glucose metabolism or insulin action leading to hyperglycemia, are already present in normoglycemia.

A novel aspect of our study is the lipidomic profiling of the PREVADIAB2, that was carried out for each cluster separated by sex and adjusted for age, BMI, and glycemic class subgroup. Lipids are a highly heterogeneous class of substances with diverse functions in the body. Recently, several works have evaluated the association of lipid species with metabolic disease (39-43), and the results have been conflicting. However, these assessments were performed in populations heterogeneous in sex and age, which may explain the incongruence of some results. Here we found that men and women displayed a distinct lipid pattern, particularly in ceramides and sphingomyelins, with higher levels of distinct circulating ceramides species (Figure 2), indicating sex differences in lipid metabolism. Women have higher SM species while men have increased CER to SM ratio. The metabolism of these sphingolipids is deeply connected, as they can be interconverted. Besides being important components of cell membrane, sphingolipids can also act as messengers (44). Sphingolipids metabolism might impact on insulin secretion and has been related with insulin resistance as well as with insulin secretion (35; 36). Indeed, it was reported that mice lacking sphingomyelin synthase 1 had insulin secretion defects and high CER to SM ratio. Interestingly, the described insulin secretion defect seemed to be mediated by ROS associated with mitochondrial dysfunction (45). Ceramide’s species have additionally been implicated in apoptosis, inflammation and have been explored and used as biomarkers for dysmetabolism and cardiovascular conditions. Particularly, β-cell apoptosis can also be partially explained by the association between ceramides and insulin deficiency.

It has been shown that circulating ceramides are originated mainly from the liver by *de novo* synthesis (42) and are higher in subjects with NAFLD. Interestingly, men appear to have a higher propensity to develop NAFLD (46).

The fact that women had different sphingolipids species than men suggests distinct CerS activities in both sexes. Specifically, women had higher dihydroceramides Cer(d16:0) and Cer(d18:0) species, particularly in the clusters with higher insulin resistance. Notwithstanding this is more evident in the dysglycemic subgroup, it was already present in women with normoglycemia. Consistently, Wigger et al. found that increased levels of dihydroceramide species Cer(d18:0/22:0) and Cer(d18:1/18:0), Cer(d18:1/20:0) and Cer(d18:1/22:0) were associated with increased susceptibility of T2D (47; 48). Particularly, they found that dihydroceramide C18:0 species were elevated in plasma sample of subjects 9 years before they developed T2D (49). However, the diagnosis of T2D in the latter study was only based in fast glucose and anti-diabetic drugs, and therefore it is not known whether the subjects already had altered postprandial glycemia. Although the results were validated by the authors in an independent cohort, it had the same drawback regarding diabetes diagnosis.

Furthermore, ceramides association with progression to T2D might be related with baseline glucose levels (50). Interestingly, despite sex differences, we found that men from PGS and IR cluster also had higher levels of Cer(d18:0/20:0) in normoglycemia or Cer(d18:0/22:0) in both normo- and dysglycemia, respectively, when compared with men from the remaining clusters. C16:0 and C18:0 dihydroceramides have been previously related to insulin resistance in other studies profile (51), even after being adjusted to BMI and visceral adipose tissue. Men had higher levels of very long-chain and ultralong chain unsaturated ceramides, namely in the clusters with low insulin levels (LS and ID), even in subjects with hyperglycemia. These species have been suggested to be related to a more metabolic healthy profile (51), associated with lower-body fat deposition, whereas LC to VLC ceramide species ratio are correlated with coronary heart disease (52). Interestingly, ultralong chain ceramides seem to be also high in women’ from PGS cluster in the subgroup with normoglycemia but not in dysglycemia subgroup.

Men additionally had a higher LPC to PC ratio. Reports regarding LPC and PC association in dysmetabolic conditions have also been conflicting. Circulating LPCs are formed from PCs, either through the action of lecithin-cholesterol acyltransferase (LCAT) or by the action of phospholipase A2 (PLA2). Therefore, LPCs levels can reflect the abovementioned enzymes activity. Indeed, LCAT activity was shown to be decreased in subjects with coronary disease (53). As for PLA2, it has been suggested that its inhibition by specific LPCs species, can explains the anti-inflammatory properties of these compounds (54). We found that LPC18:0 and LPC18:2 had higher levels in men in the LS and ID clusters with normoglycemia, decreasing in hyperglycemia. This result is consistent with other studies (55), notably in a Finnish study that included only men with normoglycemia and pre-diabetes, LPC18:2 was inversely related to progression to type 2 diabetes (13). Also, recently it has been suggested that LPC18:0 may act through PPAR gamma leading to decreased blood glucose levels (56).

The lipid signature of the clusters was consistent with their metabolic profile, showing distinct combinations between glucose and blood lipids that we postulate in conjunction with other factors are responsible for the complications of diabetes.

LS cluster showed the most metabolically favorable lipid signature, with maintained insulin clearance and higher insulin sensitivity relative to the other clusters. It showed lower levels of saturated and medium- or long-chain ceramides, as well as most TG species. Although these levels rose in the subgroup with dysglycemia, they did not appear to be higher than other clusters with apparently more lipotoxic profiles, even in cases of normoglycemia. This cluster also showed higher levels of LPC18:2 in males. This profile is consistent with a metabolically healthier cluster.

Men in the PGS cluster were all normoglycemic, and women had a low proportion of dysglycemia. Indeed, this cluster had the lowest blood glucose levels in both men and women. Furthermore, PGS was the cluster with less subjects progressing to dysglycemia. Nevertheless, this cluster showed already a lipotoxic profile in subjects with normoglycemia, which in the case of the women became more evident in the dysglycemia. Namely, saturated ceramides and TG species, such as TG48:2, which is considered a marker of *de novo* lipogenesis. Subjects from the PGS cluster had higher liver insulin resistance than those from the LS cluster but maintained peripheral insulin sensitivity. Therefore, they had the lowest glucose levels, and in men, subjects in the PGS group did not have dysglycemia. Together, results favor liver insulin resistance with increased de novo lipogenesis and, therefore, a dissociation of insulin action affection on carbohydrate and lipid metabolism in these subjects.

ID cluster in men resembled the pattern of LS clusters, even though with different levels of the lipid species. Interestingly lipid species that may be protective such as very long-chain ceramides and LPC, were present. Since it did not show hepatic insulin resistance, these results suggest that secreted insulin, although insufficient to promote glucose uptake in the periphery, has a less noticeable impact on hepatic lipid dysmetabolism and is not associated with markers of *de novo* lipogenesis.

Finally, besides glucose alterations, the IR cluster shows a lipotoxic profile, with an increase in CER and TG and a decrease in LPCs.

This work has some limitations. Namely, the results relate to an aging population, so they need to be assessed in younger populations. We could not evaluate how people evolve in these profiles over time, and we have no information to define the risk of complications in each group. We could not assess whether these patterns are: i) genetically determined, and therefore an individual trait; ii) due to lifestyle exposure and therefore depend on factors such as nutrition, exercise, or others; iii) or a mixture of the previous two. This should be evaluated in future works.

This work is not intended to define subgroups of prediabetes/diabetes. Instead, it is a proof of concept, which shows the importance of placing diabetes in a broader context of metabolism beyond glucose that considers inherent differences in age and sex as well as lipid signatures. Recently, several works have attempted to address the heterogeneity of diabetes. Diabetes is defined as hyperglycemia that results from changes in metabolism or insulin action that have many consequences other than changes in blood glucose. These changes, such as changes in lipid metabolism, contribute to the onset of diabetes complications. Interestingly, heterogeneity seems to exist before diabetes, according to the current criteria. Knowing the metabolic profiles including lipid signatures before the onset of hyperglycemia may be critical to developing more effective prevention strategies, in the context of precision medicine.

## Data Availability

All data produced in the present study are available upon reasonable request to the authors

## Acknowledgements

This work was supported by “Fundação para a Ciência e a Tecnologia” – PTDC/MEC-MET/29314/2017, 2022.05764.PTDC, UIDB/Multi/04462/2020), by the European Commission Marie Skłodowska-Curie Action H2020 (grant agreement n. 734719), by the FEDER funds through the COMPETE 2020 Program under the project BEST (ALT20-03-0247-FEDER-113469, LISBOA-01-0247-FEDER-113469) and by the Sociedade Portuguesa de Diabetologia

