## Supplementary for "Lipidomic Profiling Unveils Sex Differences in Diabetes Risk: Implications for Precision Medicine"

**Supplementary Data**

**Supplementary Figure 1.** Methodology Flowchart**.** In 2014, 1088 subjects from the PREVADIAB1 study without T2D in 2008–2009 - PREVADIAB 2 cohort. After the application of exclusion criteria and preprocessing the data, we performed two distinct cluster analyses using hierarchical clustering: one including all 953 subjects and a second analysis where individuals were separated by sex. We profiled the clusters with several metabolic parameters. Finally, we performed a lipidomic analysis of 488 individuals to further profiling the clusters, overall and separated by sex.

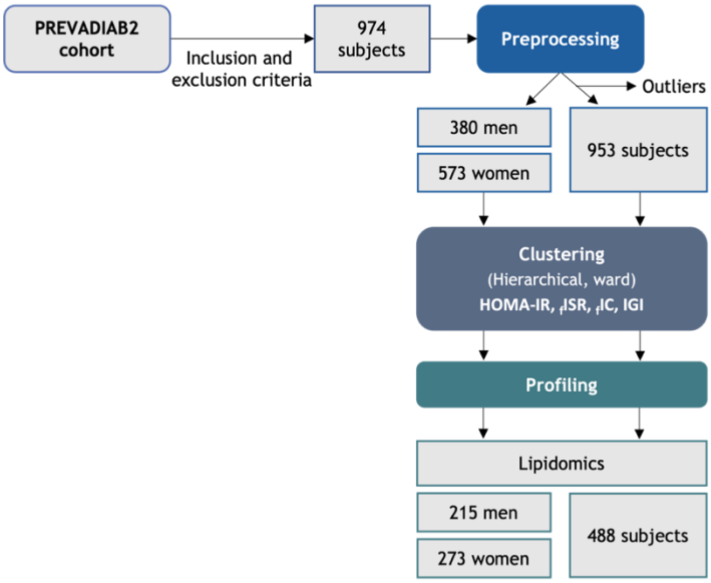

**Supplementary Figure 2.** PCA analyses of samples grouped by clusters before (A) and after (B) outliers' exclusion.

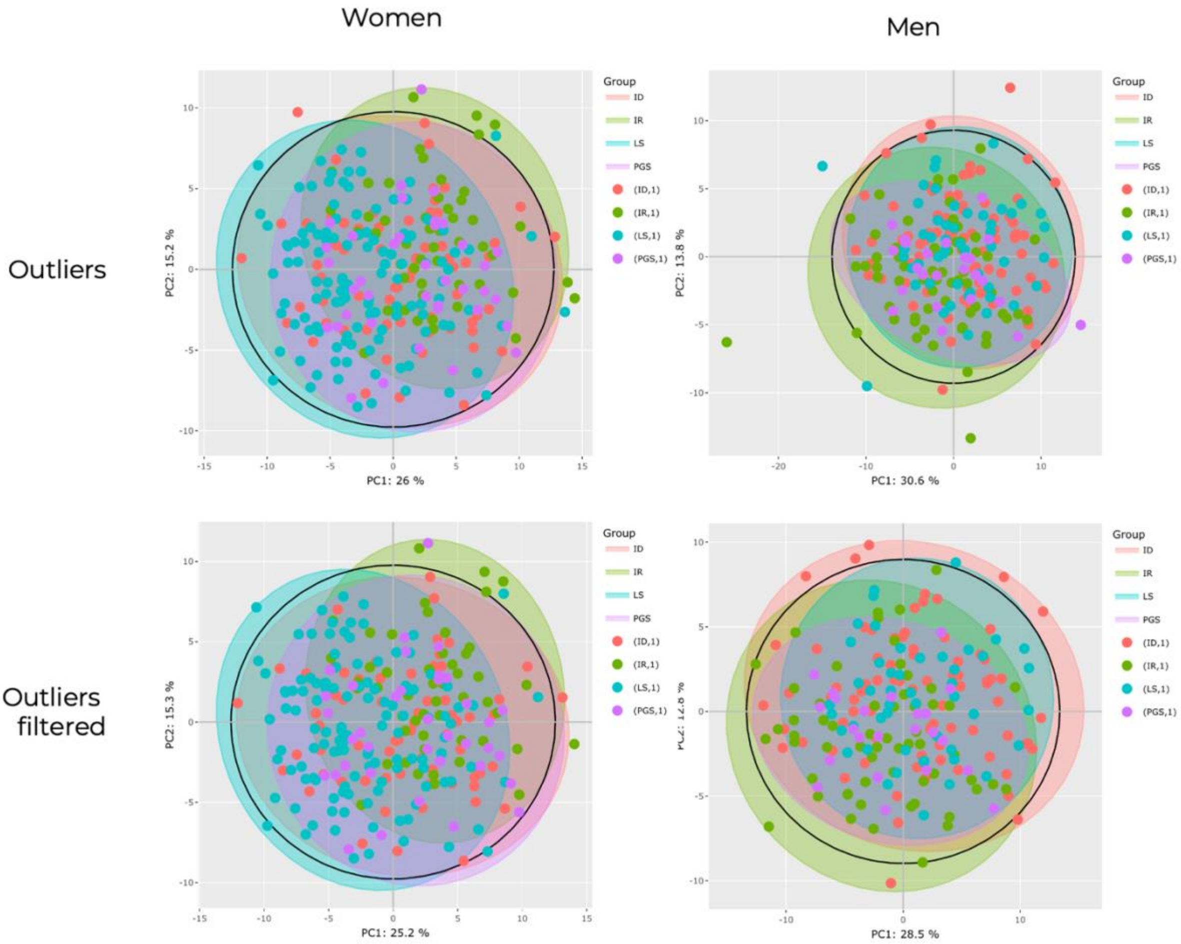

**Supplementary Figure 3. Clusters' lipidomic profiling scaled by the overall population.** Heatmap representing the scaled mean of each parameter. CER – Ceramides; ID – Insulin deficient; IR – Insulin resistant, LS – Liver sensitive; LysoPC – Lysophosphatidylcholine; PC – Phosphatidylcholine; SM – Sphingomyelin; PGS – Pancreas glucose sensitive; TG – Triglycerides.

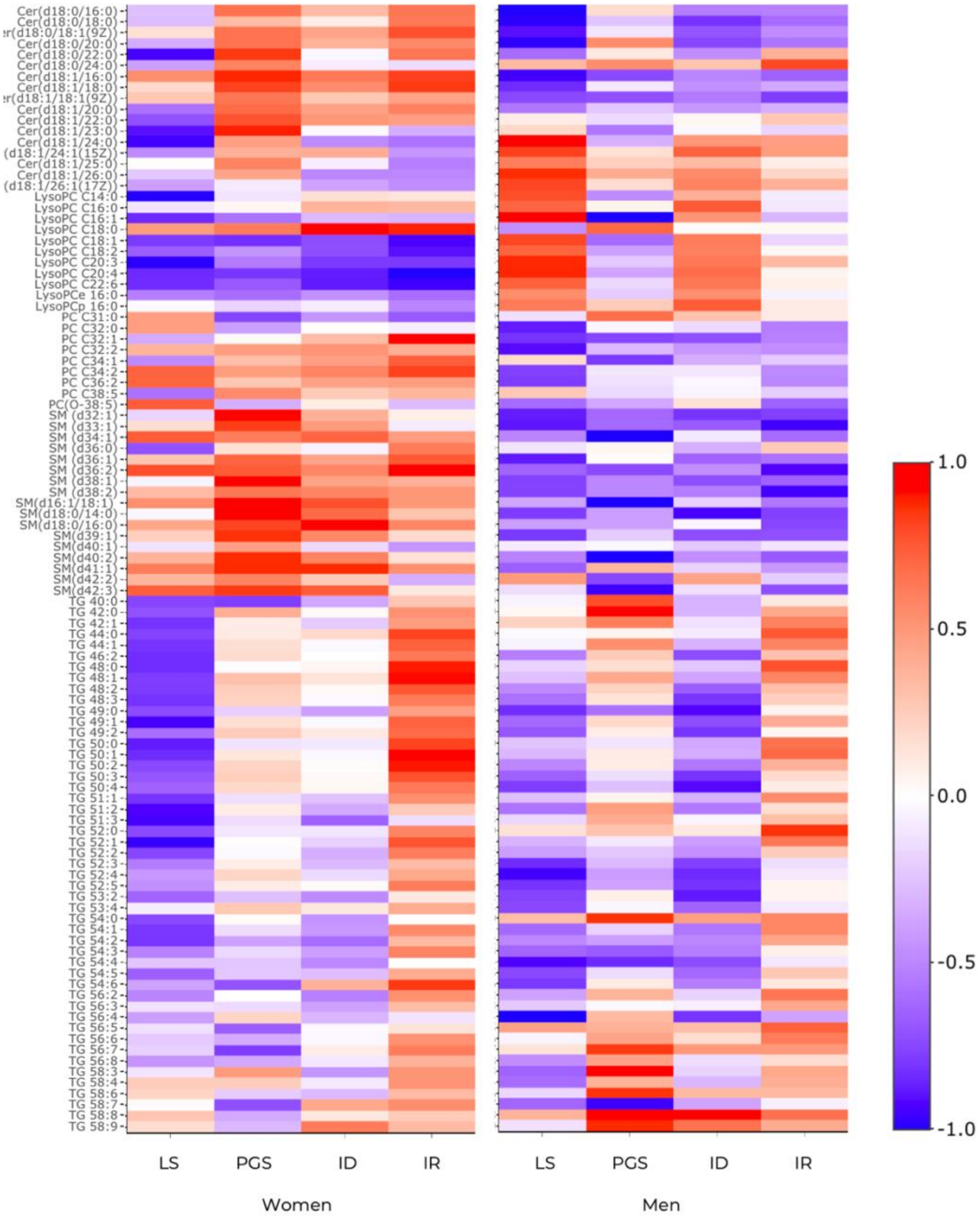

**Supplementary Figure 4. Volcano plots for lipid species-clusters association by gender.**

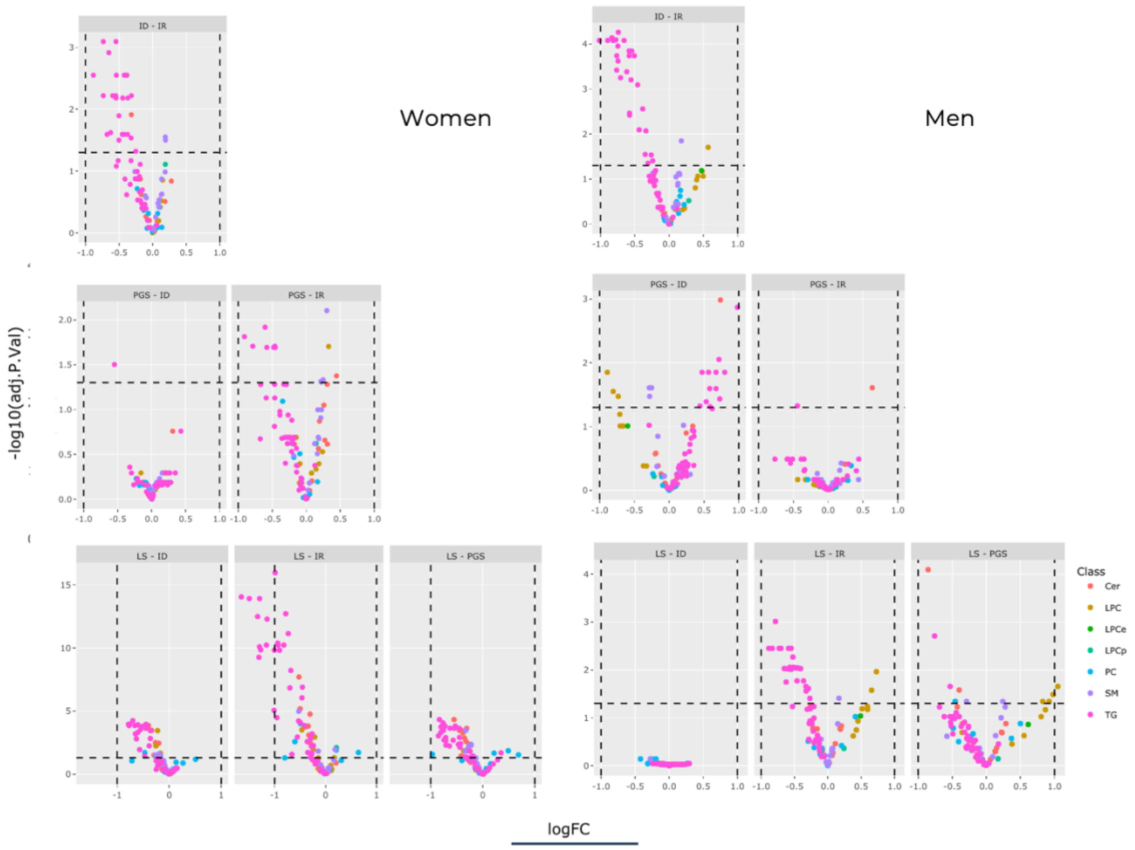

**Supplementary Figure 5. Cluster's profile by gender.** Heatmap representing the scaled mean of each parameter by gender. A) Normoglycemia subgroup; B) Dysglycemia subgroup includes prediabetes and diabetes. Cluster names: liver-sensitive (LS); pancreas glucose sensitive (PGS); insulin deficient (ID); insulin resistance (IR).

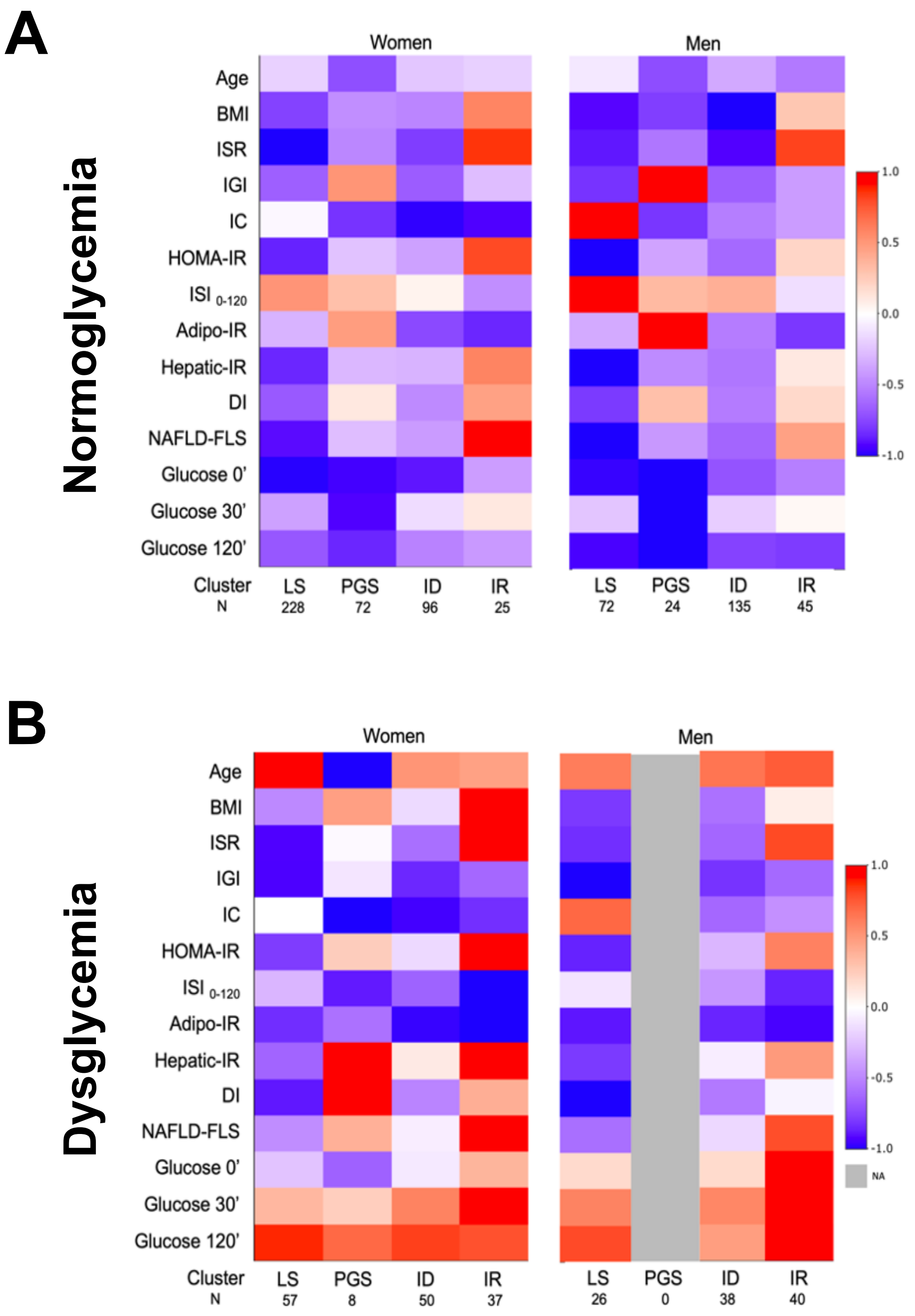

**Supplementary Table 1. Summary statistics of the population.** In bold are parameters informing cluster analysis. Values are reported as mean±SD for continuous parameters and as count and percentages for categorical parameters. Values are reported as mean±SD for continuous parameters and as count and percentages for categorical parameters. Comparison between sex was performed with Mann–Whitney test for continuous variables and with chi-square test for categorical variables.

| | All | Women | Men | P-value |
|---|---|---|---|---|
| Total n (%) | 953 | 573 (60) | 380 (40) | - |
| Normoglycemia n (%)<br>Prediabetes n (%)<br>Diabetes n (%) | 697 (73)<br>208 (22)<br>48 (5) | 421 (73)<br>125 (22)<br>27 (5) | 276 (73)<br>83 (22)<br>21 (5) | 0.1 |
| Age, Years | 61 ± 13 | 61 ± 13 | 61 ± 13 | 0.87 |
| BMI, $Kg/m^2$ | 27.3 ± 4.3 | 27.7 ± 4.6 | 26.9 ± 3.6 | 0.01 |
| **Fast-ISR, pmol/min** | **170 ± 78** | **164 ± 72** | **179 ± 84** | **0.007** |
| **IGI** | **0.90 ± 0.9** | **0.94 ± 0.9** | **0.86 ± 0.9** | **0.005** |
| **Fast-IC, L/min** | **4.2 ± 1.5** | **4.0 ± 1.4** | **4.6 ± 1.7** | **<0.001** |
| **HOMA-IR** | **1.8 ± 1.2** | **1.8 ± 1.2** | **1.8 ± 1.2** | **0.75** |
| $ISI_{0\text{-}120}$, Log10 | 67 ± 19 | 66 ± 18 | 69 ± 20 | 0.05 |
| Adipo-IR | 27 ± 19 | 27 ± 19 | 26 ± 18 | 0.5 |
| Hepatic-IR, $x10^4$ | 7.3 ± 5.4 | 7.3 ± 5.1 | 7.2 ± 5.9 | 0.05 |
| Disposition Index, $mmol^{-1}$ | 2.4 ± 2.5 | 2.4 ± 2.6 | 2.2 ± 2.3 | 0.12 |
| NAFLD-FLS | -1.2 ± 1.2 | -1.3 ± 1.2 | -1.2 ± 1.3 | 0.51 |
| Glucose 0 min, mmol/L | 5.1 ± 0.7 | 5.1 ± 0.6 | 5.3 ± 0.8 | <0.001 |
| Glucose 30 min, mmol/L | 8.6 ± 1.8 | 8.5 ± 1.7 | 8.8 ± 1.9 | 0.04 |
| Glucose 120 min, mmol/L | 6.5 ± 2.2 | 6.6 ± 2.1 | 6.3 ± 2.4 | <0.001 |

**Supplementary Table 2. Summary statistics of Clusters for the cluster analysis including the overall population.** Parameters are reported for overall population and for women and men. In bold are the parameters informing cluster analysis. Values are reported as mean±SD for continuous parameters and as count and percentages for categorical parameters. P-values are reported for comparison between sex, performed with Mann–Whitney test for continuous variables and with chi-square test for categorical variables. ISR - Insulin Secretion Rate; IC - Insulin Clearance.

| | **LS** | | | | **PGS** | | | | **ID** | | | | **IR** | | | |
|---|---|---|---|---|---|---|---|---|---|---|---|---|---|---|---|---|
| | All | Women | Men | Men vs Women (P-value) | All | Women | Men | Men vs Women (P-value) | All | Women | Men | Men vs Women (P-value) | All | Women | Men | Men vs Women (P-value) |
| Total, n (%) | 156 (16) | 285 (50) | 98 (26) | - | 78 (8) | 80 (14) | 24 (6) | - | 588 (62) | 146 (25) | 173 (46) | - | 131 (14) | 62 (11) | 85 (22) | - |
| Normoglycemia, n (%)<br>Prediabetes, n (%)<br>Diabetes, n (%) | 126 (81)<br>22 (14)<br>8 (5) | 228 (80)<br>45 (16)<br>12 (4) | 72(74)<br>21(21)<br>5(5) | 0.08* | 76 (97)<br>1 (1.5)<br>1 (1.5) | 72 (90)<br>7 (9)<br>1 (1) | 24 (100)<br>0 (0)<br>0 (0) | 0.5 | 437 (74)<br>130 (22)<br>21 (4) | 96 (66)<br>45 (31)<br>5 (3) | 135 (78)<br>33 (19)<br>5 (3) | 0.02* | 58 (44)<br>54 (41)<br>19 (15) | 25 (40)<br>28 (45)<br>9 (15) | 45 (53)<br>29 (34)<br>11 (13) | 0.6 |
| Age, Years | 62±12 | 62±13 | 62±11 | 0.8 | 55±13 | 55±13 | 56±14 | 0.8 | 61±13 | 61±12 | 60±14 | 0.6 | 62±12 | 63±12 | 62±13 | 0.7 |
| BMI, Kg/m$^2$ | 26±4 | 27±4.2 | 26±3 | 0.3 | 27±4 | 28±5 | 26±3 | 0.17 | 27±4 | 28±4 | 26±3 | <0.001* | 31±4 | 32±4 | 30±4 | <0.001* |
| **Fast-ISR, pmol/min** | 135±49 | 131±38 | 142±47 | 0.05 | 173±51 | 181±57 | 170±53 | 0.5 | 148±46 | 156±49 | 143±41 | 0.01 | 308±85 | 309±75 | 297±82 | 0.2 |
| **IGI** | 0.57±0.46 | 0.68±0.44 | 0.46±0.34 | <0.001* | 2.8±1.5 | 2.3±1.3 | 3.1±1.8 | 0.005* | 0.69±0.4 | 0.65±0.29 | 0.70±0.4 | 0.7 | 1.15±0.93 | 1.01±0.68 | 0.97±0.83 | 0.3 |
| **Fast-IC, L/min** | 6.7±1.3 | 4.9±1.1 | 6.8±1.5 | <0.001* | 3.4±0.8 | 3.3±1.1 | 3.3±0.5 | 0.1 | 3.9±1 | 2.9±0.6 | 3.8±0.9 | <0.001* | 3.4±1.1 | 3.2±0.96 | 4.1±1.0 | <0.001* |
| **HOMA-IR** | 0.74±0.28 | 1.0±0.35 | 0.81±0.34 | <0.001* | 1.91±0.70 | 2.16±0.85 | 1.9±0.7 | 0.2 | 1.51±0.63 | 1.98±0.63 | 1.55±0.72 | <0.001* | 3.9±1.06 | 4.2±1.2 | 3.2±1.2 | <0.001* |
| $ISI_{0-120}$, Log10 | 80±19 | 73±16 | 81±22 | <0.001* | 73±19 | 69±20 | 73±16 | 0.2 | 68±16 | 60±14 | 70±17 | <0.001* | 48±19 | 46±10 | 53±14 | 0.001* |
| Adipo-IR | 12±6 | 16±8 | 12±7 | <0.001* | 26±14 | 32±19 | 24±14 | 0.06 | 24±14 | 32±15 | 25±14 | <0.001* | 56±22 | 58±23 | 44±21 | <0.001* |
| Hepatic-IR, x10$^4$ | 4.3±3.5 | 5.2±2.8 | 4.3±3.9 | <0.001* | 1.2±7.1 | 1.1±6.6 | 1.2±8.2 | 0.9 | 6.2±3.3 | 6.8±3.2 | 6.4±3.6 | 0.09 | 12.9±8.0 | 12.9±7.2 | 11.0±8.2 | 0.005* |
| Disposition Index, mmol$^{-1}$ | 3.0±2.4 | 2.6±1.9 | 2.4±1.9 | 0.08 | 6.5±5.4 | 5.0±4.9 | 7±5.3 | 0.005* | 1.9±1.3 | 1.4±0.8 | 2.0±1.4 | <0.001* | 1.2±0.9 | 0.99±0.54 | 1.2±0.8 | 0.07 |
| NAFLD-FLS | -2.0±0.84 | -1.9±0.83 | -2.0±0.90 | 0.2 | -1.2±1 | -0.96±1.1 | -1.3±1.0 | 0.2 | -1.4±0.97 | -1.1±0.90 | -1.4±1.05 | <0.001* | 0.69±0.93 | 0.80±0.96 | 0.27±1.03 | 0.003* |
| Glucose 0 min, mmol/L | 4.9±0.6 | 5±0.6 | 5.1±0.7 | 0.2 | 4.8±0.4 | 4.9±0.5 | 4.9±0.4 | 0.7 | 5.1±0.6 | 5.1±0.6 | 5.2±0.6 | 0.3 | 5.7±0.8 | 5.6±0.7 | 5.7±1.0 | 0.8 |
| Glucose 30 min, mmol/L | 8.3±0.17 | 8.3±1.6 | 8.7±1.8 | 0.09 | 6.8±1.3 | 7.2±1.4 | 6.8±1.4 | 0.18 | 8.7±1.6 | 9.0±1.5 | 8.7±1.6 | 0.1 | 9.8±2.1 | 10.0±1.8 | 9.7±2.3 | 0.07 |
| Glucose 120 min, mmol/L | 6.2±2.2 | 6.3±1.9 | 6.1±2.4 | 0.06 | 5.1±1.1 | 5.6±1.6 | 4.9±1.1 | 0.05 | 6.4±2 | 7.1±2.0 | 6.1±1.9 | <0.001* | 7.7±2.8 | 7.9±2.4 | 7.3±3.1 | 0.05 |

**Supplementary Table 3.** P-values for association of lipid species with the clusters in each gender Adjusted by age, BMI and glycemic class (Normoglycemia and dysglycemia). P-values are adjusted for multicomparison (Benjamin-Hochberg). CER – Ceramides; LysoPC – Lysophosphatidylcholine; PC – Phosphatidylcholine; SM - Sphingomyelin; TG – Triglycerides.

| Species | Women | Men | Species | Women | Men |
|---|---|---|---|---|---|
| Cer(d18:0/16:0) | 3,53E-07 | 2,23E-02 | TG 40:0 | 1,44E-01 | 1,16E-02 |
| Cer(d18:0/18:0) | 5,94E-13 | 4,43E-02 | TG 42:0 | 1,15E-03 | 1,44E-04 |
| Cer(d18:0/18:1(9Z)) | 1,22E-03 | 1,18E-02 | TG 42:1 | 1,31E-04 | 2,78E-03 |
| Cer(d18:0/20:0) | 1,62E-08 | 4,76E-05 | TG 44:0 | 1,66E-09 | 6,63E-03 |
| Cer(d18:0/22:0) | 4,94E-05 | 2,08E-02 | TG 44:1 | 9,66E-09 | 6,51E-04 |
| Cer(d18:0/24:0) | 1,59E-03 | 6,05E-02 | TG 46:2 | 1,31E-10 | 4,76E-05 |
| Cer(d18:1/16:0) | 1,84E-01 | 3,96E-02 | TG 48:0 | 3,04E-14 | 1,70E-06 |
| Cer(d18:1/18:0) | 7,51E-02 | 4,57E-02 | TG 48:1 | 3,04E-14 | 4,76E-05 |
| Cer(d18:1/18:1(9Z)) | 2,91E-01 | 4,69E-01 | TG 48:2 | 2,75E-12 | 8,06E-05 |
| Cer(d18:1/20:0) | 3,04E-03 | 6,91E-02 | TG 48:3 | 4,86E-10 | 1,63E-05 |
| Cer(d18:1/22:0) | 1,29E-04 | 1,64E-02 | TG 49:0 | 5,24E-06 | 4,76E-05 |
| Cer(d18:1/23:0) | 1,39E-02 | 9,73E-02 | TG 49:1 | 1,31E-10 | 1,44E-04 |
| Cer(d18:1/24:0) | 6,17E-02 | 3,58E-02 | TG 49:2 | 4,22E-10 | 1,12E-03 |
| Cer(d18:1/24:1(15Z)) | 5,67E-02 | 1,64E-02 | TG 50:0 | 4,43E-13 | 5,68E-07 |
| Cer(d18:1/25:0) | 1,44E-03 | 1,31E-01 | TG 50:1 | 1,11E-16 | 2,47E-05 |
| Cer(d18:1/26:0) | 1,10E-03 | 1,02E-01 | TG 50:2 | 5,70E-14 | 6,51E-04 |
| Cer(d18:1/26:1(17Z)) | 1,56E-02 | 1,48E-02 | TG 50:3 | 2,75E-12 | 8,98E-04 |
| LysoPC C14:0 | 3,45E-04 | 2,55E-01 | TG 50:4 | 2,92E-09 | 2,95E-04 |
| LysoPC C16:0 | 3,69E-04 | 7,40E-01 | TG 51:1 | 1,58E-10 | 2,18E-03 |
| LysoPC C16:1 | 1,49E-02 | 1,31E-01 | TG 51:2 | 2,37E-04 | 3,94E-02 |
| LysoPC C18:0 | 4,48E-02 | 4,18E-02 | TG 51:3 | 5,67E-02 | 4,26E-01 |
| LysoPC C18:1 | 1,56E-01 | 1,00E-02 | TG 52:0 | 4,43E-13 | 1,63E-05 |
| LysoPC C18:2 | 8,21E-04 | 1,64E-02 | TG 52:1 | 3,32E-11 | 5,52E-06 |
| LysoPC C20:3 | 1,85E-01 | 2,65E-01 | TG 52:2 | 3,90E-07 | 2,74E-02 |
| LysoPC C20:4 | 8,18E-01 | 4,37E-02 | TG 52:3 | 8,67E-03 | 2,19E-01 |
| LysoPC C22:6 | 7,20E-01 | 3,32E-01 | TG 52:4 | 2,22E-02 | 9,95E-02 |
| LysoPCe 16:0 | 1,34E-01 | 1,25E-02 | TG 52:5 | 5,78E-04 | 1,60E-02 |
| LysoPCp 16:0 | 1,44E-01 | 3,10E-01 | TG 53:2 | 3,93E-02 | 8,21E-02 |
| PC C31:0 | 1,10E-03 | 2,96E-03 | TG 53:4 | 3,09E-01 | 2,99E-01 |
| PC C32:0 | 5,67E-02 | 3,03E-02 | TG 54:0 | 1,44E-04 | 4,76E-05 |
| PC C32:1 | 5,24E-06 | 4,69E-01 | TG 54:1 | 5,75E-12 | 5,68E-07 |
| PC C32:2 | 9,03E-02 | 1,02E-01 | TG 54:2 | 6,21E-07 | 1,00E-02 |
| PC C34:1 | 1,84E-02 | 9,95E-02 | TG 54:3 | 6,45E-03 | 4,98E-01 |
| PC C34:2 | 3,41E-02 | 2,42E-02 | TG 54:4 | 6,62E-01 | 5,09E-01 |
| PC C36:2 | 3,05E-01 | 1,18E-02 | TG 54:5 | 6,39E-02 | 1,28E-01 |
| PC C38:5 | 9,92E-02 | 5,10E-01 | TG 54:6 | 9,89E-05 | 9,95E-02 |
| PC(O-38:5) | 1,15E-01 | 2,55E-01 | TG 56:2 | 2,57E-05 | 1,89E-03 |
| SM (d32:1) | 2,33E-02 | 8,71E-01 | TG 56:3 | 1,00E-02 | 3,41E-01 |
| SM (d33:1) | 2,53E-02 | 4,93E-01 | TG 56:4 | 3,05E-01 | 4,35E-02 |
| SM (d34:1) | 6,26E-01 | 2,88E-01 | TG 56:5 | 2,53E-02 | 7,97E-01 |
| SM (d36:0) | 3,35E-06 | 3,03E-02 | TG 56:6 | 6,17E-02 | 6,13E-01 |
| SM (d36:1) | 6,27E-02 | 2,19E-01 | TG 56:7 | 1,37E-03 | 8,95E-01 |
| SM (d36:2) | 7,51E-02 | 4,02E-01 | TG 56:8 | 5,67E-02 | 5,84E-01 |
| SM (d38:1) | 1,07E-02 | 6,84E-01 | TG 58:3 | 1,63E-01 | 3,98E-03 |
| SM (d38:2) | 1,57E-01 | 9,95E-02 | TG 58:4 | 8,50E-01 | 6,05E-02 |
| SM(d16:1/18:1) | 2,53E-02 | 3,03E-02 | TG 58:6 | 5,24E-01 | 7,06E-01 |
| SM(d18:0/14:0) | 6,12E-02 | 6,95E-01 | TG 58:7 | 1,59E-03 | 2,23E-02 |
| SM(d18:0/16:0) | 1,84E-01 | 1,30E-01 | TG 58:8 | 1,28E-03 | 9,73E-02 |
| SM(d39:1) | 8,60E-02 | 6,45E-01 | TG 58:9 | 1,34E-01 | 6,69E-01 |
| SM(d40:1) | 8,90E-03 | 2,08E-02 | | | |
| SM(d40:2) | 2,76E-02 | 5,09E-01 | | | |
| SM(d41:1) | 1,60E-01 | 3,63E-02 | | | |
| SM(d42:2) | 3,63E-01 | 1,12E-01 | | | |
| SM(d42:3) | 1,33E-01 | 1,81E-02 | | | |

**Supplementary Table 4. Summary statistics of the clusters profiled by subgroups of normoglycemia and dysglycemia.** Values are reported as mean±sd. P-values are reported for comparison between subgroups, performed with Mann–Whitney test with Bonferroni correction. IC - Insulin Clearance; ID – Insulin deficient; IR – Insulin resistant; ISR – Insulin Secretion Rate; LS – Liver sensitive; PGS – Pancreas glucose sensitive. ns – non-significant (p>0.05)

| | LS | | | | | | PGS | | | ID | | | | | | IR | | | | | |
|---|---|---|---|---|---|---|---|---|---|---|---|---|---|---|---|---|---|---|---|---|---|
| | Women Normo | Women Dysg | P-value | Men Normo | Men Dysg | P-value | Women Normo | Women Dysg | P-value | Women Normo | Women Dysg | P-value | Men Normo | Men Dysg | P-value | Women Normo | Women Dysg | P-value | Men Normo | Men Dysg | P-value |
| N | 228 | 57 | - | 72 | 26 | - | 72 | 8 | - | 96 | 50 | - | 135 | 38 | - | 25 | 37 | - | 45 | 40 | - |
| Age, Years | 60±12 | 69±7 | <0.001 | 64±10 | 67±9 | ns | 58±13 | 54±16 | ns | 56±11 | 65±11 | 0.05 | 60±14 | 67±11 | ns | 54±16 | 65±10 | ns | 55±16 | 67±8 | 0.003 |
| BMI, m/Kg$^2$ | 27±5 | 27±4.0 | ns | 26±4 | 26±3 | ns | 27±5 | 31±3 | ns | 28±6 | 28±4 | ns | 25±2 | 27±3 | 0.003 | 34±3 | 33±4 | ns | 31±3 | 30±3 | ns |
| **Fast-ISR, pmol/min** | 135±43 | 138±35 | ns | 147±50 | 144±31 | ns | 186±58 | 220±44 | ns | 162±53 | 165±45 | ns | 132±45 | 166±39 | 0.006 | 327±96 | 313±75 | ns | 295±77 | 305±86 | ns |
| **IGI** | 0.79±0.50 | 0.37±0.26 | <0.001 | 0.58±0.43 | 0.27±0.16 | 0.02 | 2.74±1.81 | 1.54±0.71 | ns | 0.69±0.25 | 0.49±0.22 | 0.05 | 0.67±0.36 | 0.53±0.34 | ns | 1.5±0.77 | 0.81±0.44 | 0.005 | 1.15±0.89 | 0.94±1 | ns |
| **Fast-IC, Lmin$^{-1}$** | 4.9±1.0 | 5.0±1.2 | ns | 6.5±1.0 | 6.4±1.0 | ns | 3.5±1.0 | 2.9±1.1 | ns | 3.1±0.5 | 3.0±0.6 | ns | 4.1±0.8 | 3.8±0.8 | ns | 3.1±0.9 | 3.2±1.0 | ns | 4.1±1.0 | 4.1±1.1 | ns |
| **HOMA-IR** | 1.0±0.39 | 1.1±0.36 | ns | 0.85±0.32 | 0.95±0.31 | ns | 2.1±0.86 | 2.9±0.61 | ns | 2.0±0.7 | 2.2±0.58 | ns | 1.24±0.5 | 1.9±0.79 | <0.001 | 4.3±1.33 | 4.3±1.16 | ns | 2.91±1.04 | 3.57±1.20 | ns |
| $ISI_{0-120}$, Log10 | 77±15 | 58±7 | <0.001 | 89±21 | 63±16 | <0.001 | 71±21 | 43±5 | <0.001 | 62±14 | 49±9 | <0.001 | 78±18 | 53±9 | <0.001 | 52±13 | 40±6 | 0.005 | 60±16 | 45±8 | <0.001 |
| Adipo-IR | 16±9 | 21±10 | 0.05 | 13±8 | 16±7 | ns | 28±17 | 62±14 | <0.001 | 30±17 | 39±16 | ns | 19±10 | 35±14 | <0.001 | 55±27 | 63±21 | ns | 42±27 | 49±21 | ns |
| Hepatic-IR, *10$^4$ | 5.6±2.9 | 4.0±2.1 | 0.04 | 4.7±4.2 | 3.3±2.0 | ns | 11.8±7.7 | 16.8±3.0 | ns | 7.6±3.6 | 6.6±3.6 | ns | 5.5±3.4 | 6.3±3.7 | ns | 15.7±9.5 | 13.0±6.9 | ns | 12.8±7.8 | 10.9±9.8 | ns |
| Disposition Index, mmol$^{-1}$ | 2.9±1.9 | 1.3±0.9 | <0.001 | 2.6±1.8 | 1.2±0.6 | 0.02 | 6.1±6.2 | 2.1±1.3 | 0.01 | 1.5±0.9 | 0.9±0.5 | ns | 2.3±1.3 | 1.3±0.9 | <0.001 | 1.3±0.5 | 0.8±0.3 | 0.003 | 1.5±0.7 | 1.1±1.0 | ns |
| NAFLD-FLS | -2.0±0.81 | -1.4±0.81 | 0.006 | -2.0±0.73 | -1.5±0.84 | ns | -1.1±0.96 | -0.1±1.2 | ns | -1.2±0.96 | -0.8±0.83 | ns | -1.8±0.98 | -1.0±1.01 | 0.005 | 0.96±1.031 | 0.88±0.98 | ns | 0.27±1.2 | 0.53±1.01 | ns |
| Glucose 0' min, mmol/L | 4.9±0.48 | 5.4±0.7 | <0.001 | 4.9±0.55 | 5.7±0.84 | <0.001 | 4.9±0.47 | 5.1±0.61 | ns | 4.9±0.51 | 5.5±0.73 | <0.001 | 5.1±0.45 | 5.7±0.85 | 0.002 | 5.3±0.51 | 5.8±0.76 | 0.08 | 5.2±0.52 | 6.3±0.98 | <0.001 |
| Glucose 30' min, mmol/L | 8.0±1.46 | 9.5±1.38 | <0.001 | 8.3±1.47 | 9.9±2.0 | 0.002 | 7.0±1.29 | 9.23±1.24 | 0.003 | 8.5±1.35 | 9.9±1.41 | <0.001 | 8.4±1.36 | 9.9±1.91 | <0.001 | 9.0±1.61 | 10.7±1.50 | <0.001 | 8.8±1.53 | 10.6±2.66 | 0.03 |
| Glucose 120' min, mmol/L | 5.6±1.16 | 9.3±1.57 | <0.001 | 5.0±1.31 | 9.1±2.24 | <0.001 | 5.2±1.07 | 8.9±1.60 | <0.001 | 6.0±1.13 | 9.2±1.72 | <0.001 | 5.4±1.20 | 8.3±2.18 | <0.001 | 6.2±1.0 | 9.1±2.45 | <0.001 | 5.4±1.13 | 9.6±3.0 | <0.001 |
